# An updated method to estimate factors associated with tuberculosis transmission using whole genome sequencing and other additional metadata

**DOI:** 10.64898/2025.12.09.25341923

**Authors:** Anne N. Shapiro, ChuanChin Huang, Meredith B. Brooks, Samantha Malatesta, Leonid Lecca, Mercedes C. Becerra, Roger I. Calderon, Carmen C. Contreras, Judith Jimenez, Rosa Yataco, Zibiao Zhang, Megan B. Murray, Laura F. White, Helen E. Jenkins

## Abstract

**Background:** Understanding tuberculosis (TB) transmission dynamics is necessary to interrupt disease spread. We developed a model to estimate adjusted odds ratios (ORs) for factors associated with genetic relatedness, as proxy for transmission.

**Methods:** We build upon an existing iterative model that modifies genetically linked tuberculosis case data to better represent true transmission links. We incorporate bootstrapped logistic regression to calculate adjusted ORs with confidence intervals that account for correlation from individuals present across multiple transmission pairs. We assess model performance with simulation studies and apply the method to cohort data from Lima, Peru.

**Results:** Iterative algorithm estimates resembled those from logistic regression but had larger confidence intervals, reflecting the data correlation adjustment. Transmission pairs where at least one member was >34 years had decreased transmission odds. Pairs with at least one incarcerated or male member had increased adjusted transmission odds.

**Conclusions:** We produce adjusted ORs accounting for the correlation of pairwise genetic relatedness data. These ORs are an accurate proxy for the association between covariates and transmission and further our understanding of factors associated with tuberculosis transmission.

## 1. INTRODUCTION

Tuberculosis (TB) is the leading cause of death from a single infectious agent globally, infecting and killing millions annually despite global efforts to combat TB.^1^ Challenges in interrupting TB transmission include disease burden heterogeneity ^2, 3^ and slow disease progression, such that transmission events are often unobservable and difficult to trace.^4^ In this data-limited setting, modeling can serve as a tool to help infer disease dynamics. Understanding TB transmission dynamics and risk factors is important for informing targeted interventions to reduce the spread of, and ultimately eliminate, TB.

Multiple studies estimate transmission risk factors using various methods.^5, 6, 7, 8^ Most rely on genetic differences between the infecting *Mycobacterium tuberculosis* (M.tb), which cannot definitively determine transmission^9^ and/or modeling phylogenetic trees, which requires strong assumptions^10, 11, 12, 13^ A recent study by Trevisi et al.^14^ used multivariable logistic regression on pairs of people with TB to estimate odds ratios (ORs) of factors associated with TB transmission, as defined by single nucleotide polymorphisms (SNP) distances between the infecting M.tb obtained from whole genome sequencing (WGS). However, concerns were raised that not accounting for correlation present in the pairwise genetic data used in the analysis resulted in artificially small standard errors and that a single SNP cutoff cannot determine transmission.^15, 16^

In response, we build upon previous work^17^ (henceforth “mlTransEpi”) to redo the analysis presented in Trevisi et al.^14^ mlTransEpi is an iterative framework to calculate transmission probabilities between two individuals in a TB outbreak and univariate (unadjusted) ORs of the association between covariates and genetic relatedness (as defined by SNP distances between the infecting M.tb), which we showed to be adequate proxies for the association between covariates and transmission.^17^ We expand mlTransEpi to calculate adjusted ORs using a bootstrapped logistic regression, which provides standard errors robust to correlation.^18^ Adjusted ORs are important because TB exhibits significant disease burden heterogeneity and many known risk factors impact transmission rates across demographics.^19, 20^ Understanding how these risk factors work together in an adjusted model, as Trevisi et al. did,^14^ is necessary for accurate transmission risk estimation. A strength of mlTransEpi is that it does not require the strong assumptions or high computational costs necessary in generating phylogenetic trees.^9^

This paper extends the mlTransEpi framework to estimate adjusted ORs of associations between covariates and genetic relatedness, serving as a transmission proxy, and use it to re-estimate the ORs produced in Trevisi et al.^14^ We calculate adjusted ORs via a bootstrapped logistic regression model nested within mlTransEpi’s iterative model, which modifies the pairwise genetic relatedness data to more closely resemble transmission data and accounts for correlation present from the pairwise data structure. We show via simulation studies that these adjusted ORs capture the covariate-transmission associations.

## 2. METHODS

### 2.1 Naïve Bayes Transmission Method Odds Ratios

To understand TB transmission dynamics, ideally we would estimate the OR for transmission and covariates (OR^T^). However, this is unfeasible as transmission events are typically unobservable. As a proxy, we can estimate the OR describing the association between genetic relatedness and transmission (OR^G^). We expect OR^G^ to be similar but not identical to OR^T^, as genetic relatedness does not necessarily imply direct transmission.

In mlTransEpi, we improved upon estimates of OR^G^ through an iterative estimation procedure that modifies the dataset of genetic links to better resemble the dataset of true (unobserved) transmission links, and then used this dataset to produce new estimates that we will refer to as OR^M^.^17^ Figure 1 shows the relationship between OR^T^, OR^G^, and OR^M^ and the iterative procedure is shown in Figure 2 and detailed in Supplementary Materials Section S1 and elsewhere.^17, 21^ Using genetic relatedness to inform probable transmission links as in OR^G^ may result in one case having probable links with multiple infectors, despite there only being one true infector. The iterative procedure corrects for this in OR^M^ by randomly choosing one true infector in each iteration amongst all possible infectors (based on SNP distances) and designating all other possible infectors as non-transmission links. Multiple iterations ensure each possible infector is considered. OR^M^ estimates are averaged across all iterations, with variances pooled using Rubin’s rules,^22^ to achieve a final estimate.

**Figure 1.**
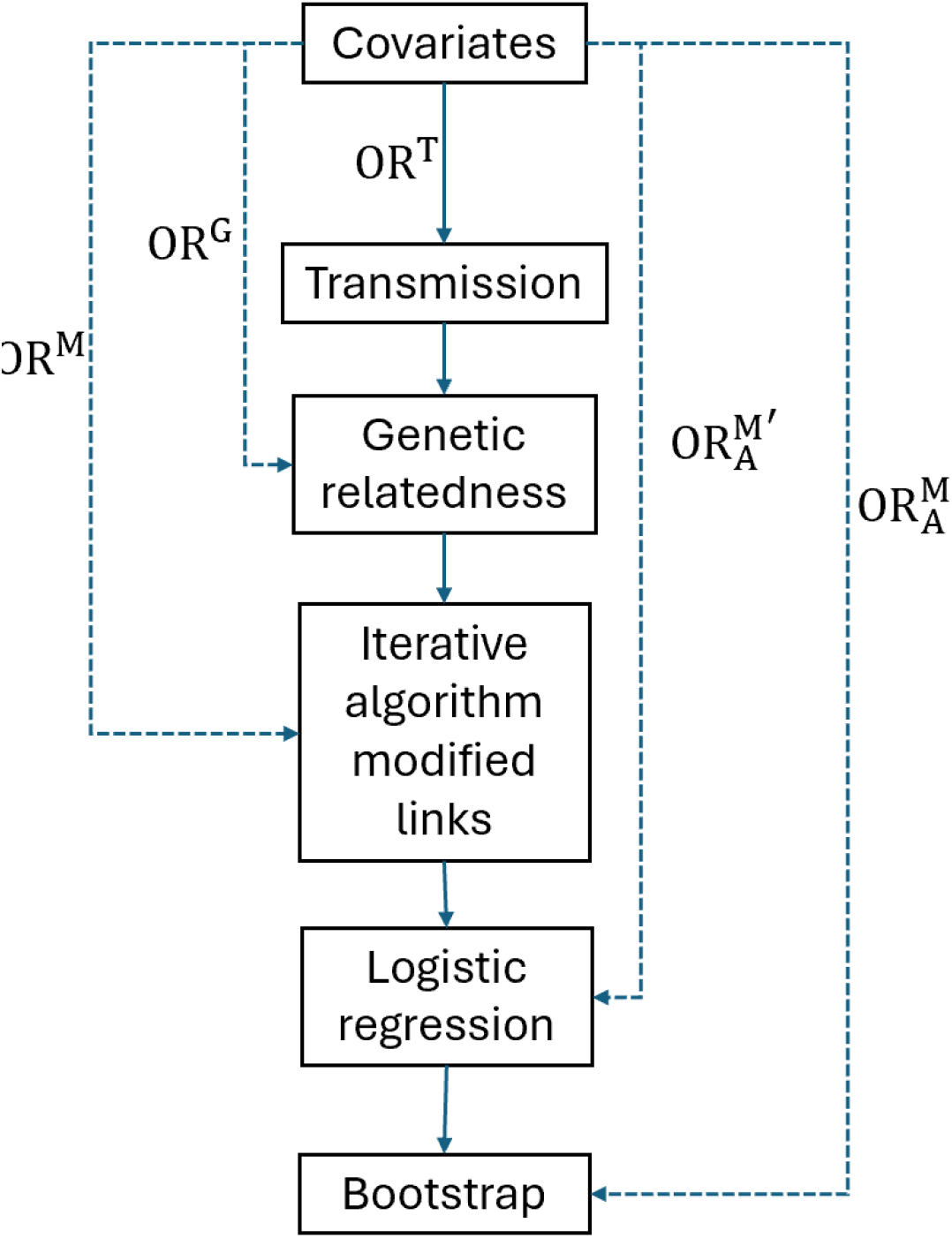
Diagram of the relationships of the odds ratios described and calculated in this paper. The unadjusted association between covariates and transmission, which is usually unobserved, is represented as OR^T^ and the relationship between covariates and observed genetic relatedness assess by single nucleotide polymorphisms (SNP) distances (as a proxy for transmission) is represented as OR^G^. OR^M^ refers to the odds ratio between covariates and genetic links modified through the iterative algorithm. We also generate non-bootstrapped adjusted associations between covariates links modified through the iterative algorithm via logistic regression 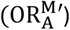, and bootstrapped adjusted logistic regression odds ratios (to account for correlation) within the iterative using modified links 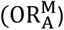.

**Figure 2.**
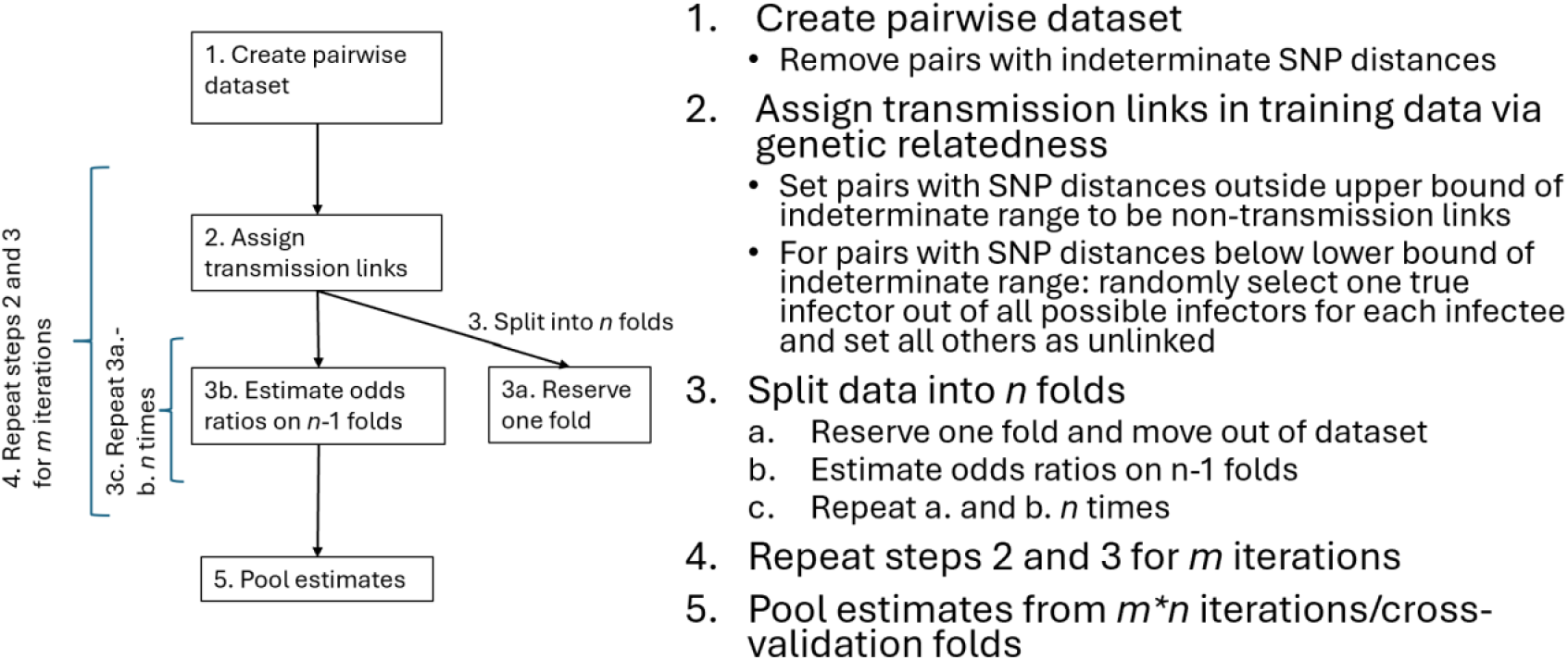
Diagram of the iterative process to generate odds ratios.

### 2.2 Adjusted Odds Ratios

We extend mlTransEpi to calculate adjusted ORs that account for the correlation induced by using a pairwise dataset through (1) using the dataset modified to better resemble true transmission links such that only one true infector exists for each infectee and (2) bootstrapping the adjusted OR to calculate non-parametric errors that are robust to correlation.^18^ We will refer to this estimate as 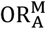. Note all ORs described in Section 2.1 are unadjusted. Figure 2 shows the relationship between 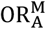, OR^T^, OR^G^ and OR^M^.

For a model run with *m* iterations and *n* within-iteration folds, the iterative estimation procedure results in *m* ∗ *n* estimates of each 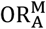, for each of which we estimate and pool 100 *m* bootstrap samples. Further bootstrap details are presented in Supplementary Materials Section 2. The final estimate of is calculated by averaging 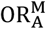 across all iterations; standard errors are pooled using Rubin’s rules.^22^

### 2.3 Simulation study

We simulate 500 outbreaks using R v4.4.0 informed by TB epidemiology. ^17, 21^ For each outbreak we simulate six categorical pairwise covariates (*Z*_*i*_, *i* = 1, …, 6) using the transmission tree structure, such all covariates are associated with transmission, and simulate covariates *Z*_1_ and *Z*_5_ to be correlated. Simulation structure is described in detail Supplementary Materials Section 2.

We run each outbreak through mlTransEpi with 10 iterations and 10 within-iteration folds. Pairs with <4 SNP differences are considered probable transmission links; pairs with >12 SNP differences are probable non-links. Pairs with 4-12 SNP differences are considered indeterminate and excluded from the training dataset. For each outbreak, we calculate OR^T^, OR^G^, OR^M^, 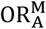, and an adjusted OR from logistic regression without bootstrapping 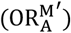 for each outbreak. 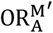 does not account for correlation, so its standard errors are expected to be underestimates (i.e. smaller than those of 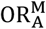). Figure 2 shows how 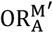 relates to other ORs.

We calculate the mean absolute percentage error (MAPE) and mean squared error (MSE) with respect to OR^T^ (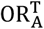, an adjusted OR describing the association between covariates and transmission) for OR^G^ and OR^M^ (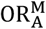 and 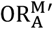). We calculate confidence interval (CI) coverage as the percentage of time that 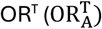 is within the CI of OR^G^ and OR^M^ (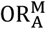 and 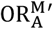), and the CI width for all estimates.

We perform sensitivity analyses on simulation assumptions. We examine using multiple lower SNP distance lower thresholds, scale parameters of the gamma distributions describing the generation interval and sampling distribution, and reproductive numbers. We also vary the proportion of the outbreak sampled and the time allowed between potential infectors and infectees. For each setting, we estimate and compare values of OR^T^, OR^M^, 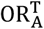, and 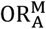.

Functions to calculate OR^M^ and 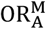 are implemented in the R package nbTransmission.^21^

### 2.4 Data application

The original study by Trevisi et al. identified factors associated with TB transmission in a large prospective cohort in Lima, Peru.^14^ The data consist of individuals aged 16 or older with pulmonary TB diagnosed at a participating health center.^23^ Trevisi et al. assigned pairs as direct transmission (< 4 SNP differences) or non-transmission (>=4 SNP differences). They created pairwise covariates and used logistic regression to calculate OR based on SNP-defined transmission,^14^ assuming pair independence.^24^

We apply mlTransEpi to data from Trevisi et al.^14^ with 60 iterations and 10 within-iteration folds. We used 60 iterations to ensure each of the up to 32 potential infectors per individual had ample opportunity to be selected as the ‘true’ infector. Pairs with <4 SNP differences are probable links, >12 SNP differences are probable non-links, and 4-12 SNP differences are indeterminate. We considered pairs where an infector was observed more than 60 days before an infectee, as in Trevisi et al.^14^ Pairwise covariates were created as described in the original paper.

Due to the size of the dataset (>3 million pairs), a full bootstrap within the iterative framework is computationally intensive. Instead, we sample all transmission pairs (SNP distances < 4; n = 2,481) and 100,000 non-transmission pairs (SNP distances > 12; n = 3,069,784) with replacement for 100 bootstrap replicates. We assess via sensitivity analysis within a single logistic regression that OR estimates and standard errors from samples of this size were consistent with OR estimates and standard errors from a traditional bootstrap after an adequate sample size was reached (Supplemental Materials Figure S2).

We first calculate adjusted OR using the same variables included in Trevisi et al.’s multivariable model^14^ to directly compare mlTransEpi estimates 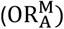 the logistic regression estimates (OR_A_). We then repeat the model selection performed in Trevisi et al.^14^ using unadjusted ORs from mlTransEpi. Categorical variables were defined as in their work. We consider people public transit users if they use it ≥4 times/week. A socioeconomic status (SES) score, created via principal component analysis, was dichotomized into low (first tercile) and not low (second/third terciles). Smokers and drinkers consumed ≥1 cigarette or alcoholic drink/day, respectively. Univariate OR (OR^M^) were calculated for all variables described in Table 1. Covariates with at least one OR^M^ p-value ≤ 0.1 were included in a multivariable model for 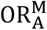. This analysis included all pairs where the potential infector was observed prior to the potential infectee.

**Table 1.** Cohort pair-level demographic and clinical characteristics, n (%) stratified by single nucleotide polymorphisms (SNP) differences.

| Variable | Level | Total pairs<br>(n = 3,078,413) | Pairs with SNP<br>Differences < 4<br>(n = 2,481) | Pairs with SNP<br>Differences > 12<br>(n = 3,069,784) |
| --- | --- | --- | --- | --- |
| Age (years) | >34 → >34 | 719,262 (23.36) | 445 (17.94) | 717,424 (23.37) |
|  | >34 → ≤ 34 | 684,504 (22.24) | 510 (20.56) | 682,754 (22.24) |
|  | ≤ 34 → >34 | 1,295,056 (42.07) | 1,345 (54.21) | 1,290,844 (42.05) |
|  | ≤ 34 → ≤ 34 | 465,730 (15.13) | 268 (10.8) | 464,830 (15.14) |
| Sex | F → F | 734,742 (23.87) | 523 (21.08) | 732,934 (23.88) |
|  | F → M | 729,298 (23.69) | 574 (23.14) | 727,331 (23.69) |
|  | M → F | 1,148,643 (37.31) | 1,116 (44.98) | 1,144,689 (37.29) |
|  | M → M | 2,111,652 (68.6) | 1,494 (60.22) | 2,106,210 (68.61) |
| Work outside<br>the home | No → No | 292,440 (9.5) | 298 (12.01) | 291,545 (9.5) |
|  | No → Yes | 216,035 (7.02) | 228 (9.19) | 215,330 (7.01) |
|  | Yes → No | 262,829 (8.54) | 264 (10.64) | 261,930 (8.53) |
|  | Yes → Yes | 195,457 (6.35) | 197 (7.94) | 194,769 (6.34) |
| Public transit<br>use | Heavy → Light | 709,513 (23.05) | 586 (23.62) | 707,571 (23.05) |
|  | Light → Heavy | 736,082 (23.91) | 573 (23.1) | 733,940 (23.91) |
|  | Light → Light | 435,319 (14.14) | 305 (12.29) | 434,083 (14.14) |
| Socioeconomic<br>status | Low → Low | 646,099 (20.99) | 523 (21.08) | 644,222 (20.99) |
|  | Low → Not low | 750,293 (24.37) | 644 (25.96) | 748,142 (24.37) |
|  | Not low → Low | 778,751 (25.3) | 617 (24.87) | 776,572 (25.3) |
|  | Not low → Not low | 903,270 (29.34) | 697 (28.09) | 900,848 (29.35) |
| Spent at least<br>one night in<br>prison | No → No | 335,815 (10.91) | 323 (13.02) | 334,670 (10.9) |
|  | No → Yes | 722,637 (23.47) | 541 (21.81) | 720,580 (23.47) |
|  | Yes → No | 639,944 (20.79) | 510 (20.56) | 638,055 (20.79) |
|  | Yes → Yes | 1,380,017 (44.83) | 1,107 (44.62) | 1,376,479 (44.84) |
| Smoking status | Nonsmoker → Nonsmoker | 2,797,980 (90.89) | 1,806 (72.79) | 2,791,097 (90.92) |
|  | Nonsmoker → Smoker | 135,689 (4.41) | 255 (10.28) | 134,945 (4.4) |
|  | Smoker → Nonsmoker | 138,115 (4.49) | 343 (13.83) | 137,258 (4.47) |
|  | Smoker → Smoker | 6,629 (0.22) | 77 (3.1) | 6,484 (0.21) |
| Drinking status | Nondrinker → Nondrinker | 2,922,026 (94.92) | 2,295 (92.5) | 2,914,019 (94.93) |
|  | Nondrinker → Drinker | 75,012 (2.44) | 63 (2.54) | 74,758 (2.44) |
|  | Drinker → Nondrinker | 79,375 (2.58) | 114 (4.59) | 79,024 (2.57) |
|  | Drinker → Drinker | 2,000 (0.06) | 9 (0.36) | 1,983 (0.06) |
| HIV status | Negative → Negative | 985,536 (32.01) | 572 (23.06) | 983,460 (32.04) |
|  | Negative → Positive | 699,378 (22.72) | 520 (20.96) | 697,400 (22.72) |
|  | Positive → Negative | 813,718 (26.43) | 727 (29.3) | 811,375 (26.43) |
|  | Positive → Positive | 579,781 (18.83) | 662 (26.68) | 577,549 (18.81) |
| Diabetes | No → No | 2,866,811 (93.13) | 2,320 (93.51) | 2,858,745 (93.13) |
|  | No → Yes | 98,291 (3.19) | 69 (2.78) | 98,032 (3.19) |
|  | Yes → Everyone else | 109,590 (3.56) | 86 (3.47) | 109,299 (3.56) |
| Cavity | No → Everyone else | 3,721 (0.12) | 6 (0.24) | 3,708 (0.12) |
|  | Yes → Everyone else | 2,743,900 (89.13) | 2,290 (92.3) | 2,735,969 (89.13) |
| Cough | No → Everyone else | 162,351 (5.27) | 90 (3.63) | 161,967 (5.28) |
|  | Yes → Everyone else | 172,162 (5.59) | 101 (4.07) | 171,848 (5.6) |
| Prior TB history | No → Everyone else | 2,211,442 (71.84) | 1,742 (70.21) | 2,205,274 (71.84) |
|  | Yes → Everyone else | 866,971 (28.16) | 739 (29.79) | 864,510 (28.16) |
| Smear | Negative → Everyone else | 1,520,228 (49.38) | 1,265 (50.99) | 1,516,201 (49.39) |
|  | Positive → Everyone else | 1,558,185 (50.62) | 1,216 (49.01) | 1,553,583 (50.61) |

## 3. RESULTS

### 3.1 Simulation study

Results from the simulation study are shown in Figure 3. Recall that covariates *Z*_1_ and *Z*_5_were simulated to be correlated. In the unadjusted analyses describing the association between covariates and transmission (OR^T^), all covariates except *Z*_3_ are strongly associated with transmission. After adjusting, both *Z*_1_ and *Z*_5_ become much closer to the null. OR^M^ and 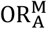, the unadjusted and adjusted iteratively modified OR describing the association between transmission and covariates, follow similar trends but are all biased towards the null compared to OR^T^ and 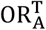, respectively. Results show the same patterns across varying the reproductive number and scale parameters of the generation interval and sampling distribution (Supplemental Materials Figure S3). As the SNP distance threshold increases, all estimates move slightly closer to the null (Supplemental Materials Figure S4).

**Figure 3.**
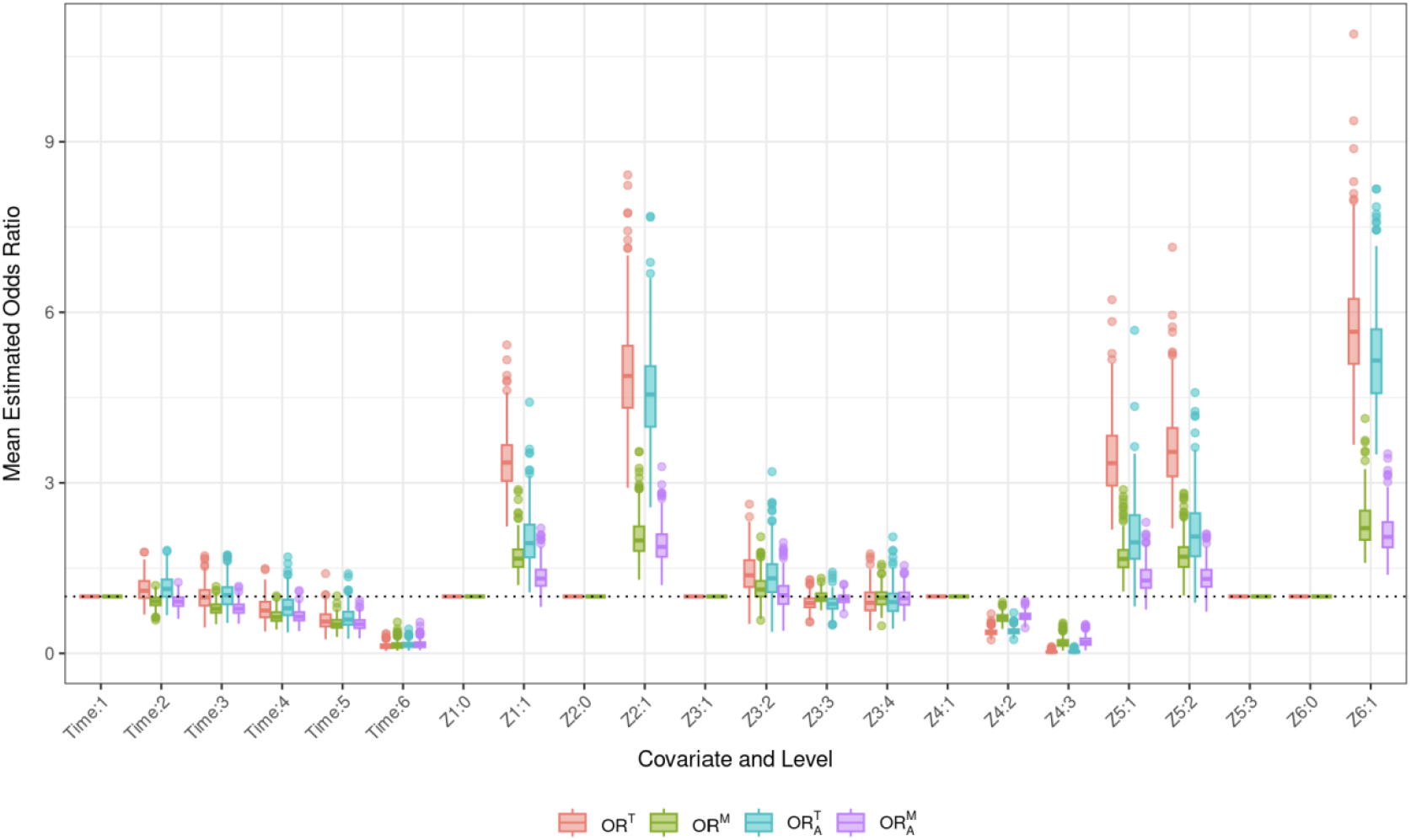
Mean estimated odds ratios from 500 simulated outbreaks describing the unadjusted association between covariates and transmission (OR^T^), adjusted association between covariates and transmission 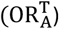, unadjusted odds ratios describing the association between covariates and iteratively modified close genetic relatedness (OR^M^), and adjusted bootstrapped odds ratios describing the association between covariates and iteratively modified close genetic relatedness 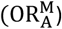. Covariates Z1 and Z5 are simulated to be correlated. We are interested in obtaining OR^T^ and 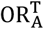 but these quantities are not measurable so we estimate OR^M^ and 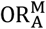 as a proxy.

MSE and MAPE are lower for 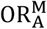, and 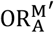 compared to OR^M^, indicating that they more closely capture 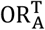 than OR^M^ captures OR^T^ (Supplemental Materials Figure S5).

### 3.2 Data application

Covariates stratified by probable links (SNP distance < 4) and non-links (SNP distance > 12) are shown in Table 2.

Adjusted ORs from the iterative method (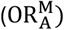) and original paper (OR_A_) are presented in Figure 4. As expected, point estimates from both models are similar but CIs generated by the iterative model are often wider, reflecting the adjustment for correlation in the data. Transmission pairs with the infector ≤34 years old and the infectee > 34 years old have significantly lower 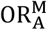 compared to pairs with both the infector and infectee ≤34 years old. Transmission pairs with a male infector and female infectee and both infector and infectee being smokers no longer have significantly increased 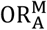 compared to pairs with female infectors and infectees and non-smoking infectors and infectees, respectively.

**Figure 4.**
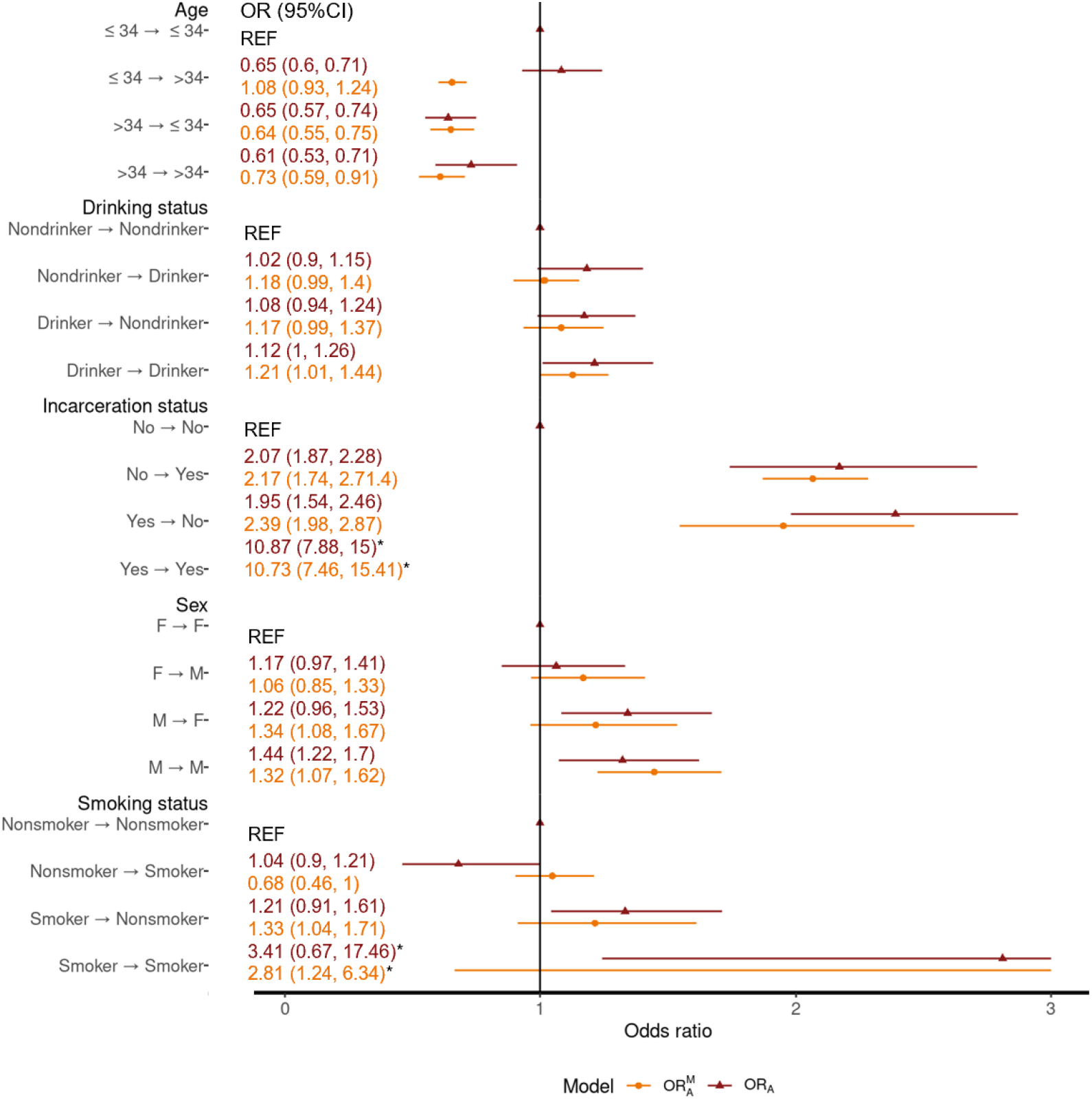
Estimated adjusted naïve Bayes modified close genetic relatedness odds ratios 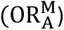 and odds ratios estimated by a multivariable logistic regression model (OR_A_) for the Lima, Peru cohort dataset. Both estimates are presented with 95% confidence intervals. *The value is out of scale and has not been included or entirely represented.

Unadjusted OR^M^ are presented in Supplemental Materials Table S2. Variables describing age, sex, drinking status, incarceration status, smoking status, prior TB history, socioeconomic status, and time between diagnosis were found to be significant at the 0.1 level and included in the multivariable model. In the multivariable model, transmission pairs consisting of someone > 34 had significantly lower 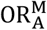 compared to pairs with both individuals being ≤34 (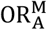 (95% CI) ≤ 34 → >34: 0.65, (0.59, 0.71); >34 → ≤ 34: 0.64 (0.55, 0.73); >34 → >34: 0.61 (0.52, 0.7)) (Figure 5). Pairs with individuals both considered drinkers and both males had increased odds of transmission (drinker → drinker 1.14 (1.02, 1.27); M → M 1.42 (1.22, 1.65)) compared to pairs with both individuals considered nondrinkers and females, respectively. All transmission pairs consisting of one individual not in the lowest socioeconomic status (SES) group (bottom 33% of adjusted SES score) had significantly lower 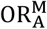 compared to pairs with both individuals in the lowest SES group (Low → Not low: 0.65(0.55, 0.78); Not low → Low: 0.74 (0.61, 0.9); Not low → Not low: 0.83 (0.73, 0.95)). Pairs in which the infector had a previous TB diagnosis had 17% increased odds (95% CI (1.02, 1.34)) of being a transmission pair. 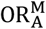 for time between diagnosis decreased as the time increased (3-6 months: 0.76 (0.65, 0.88); 6-12 months: 0.65 (0.56, 0.75); 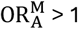 year: 0.5 (0.41, 0.62) (reference <3 months)).

**Figure 5.**
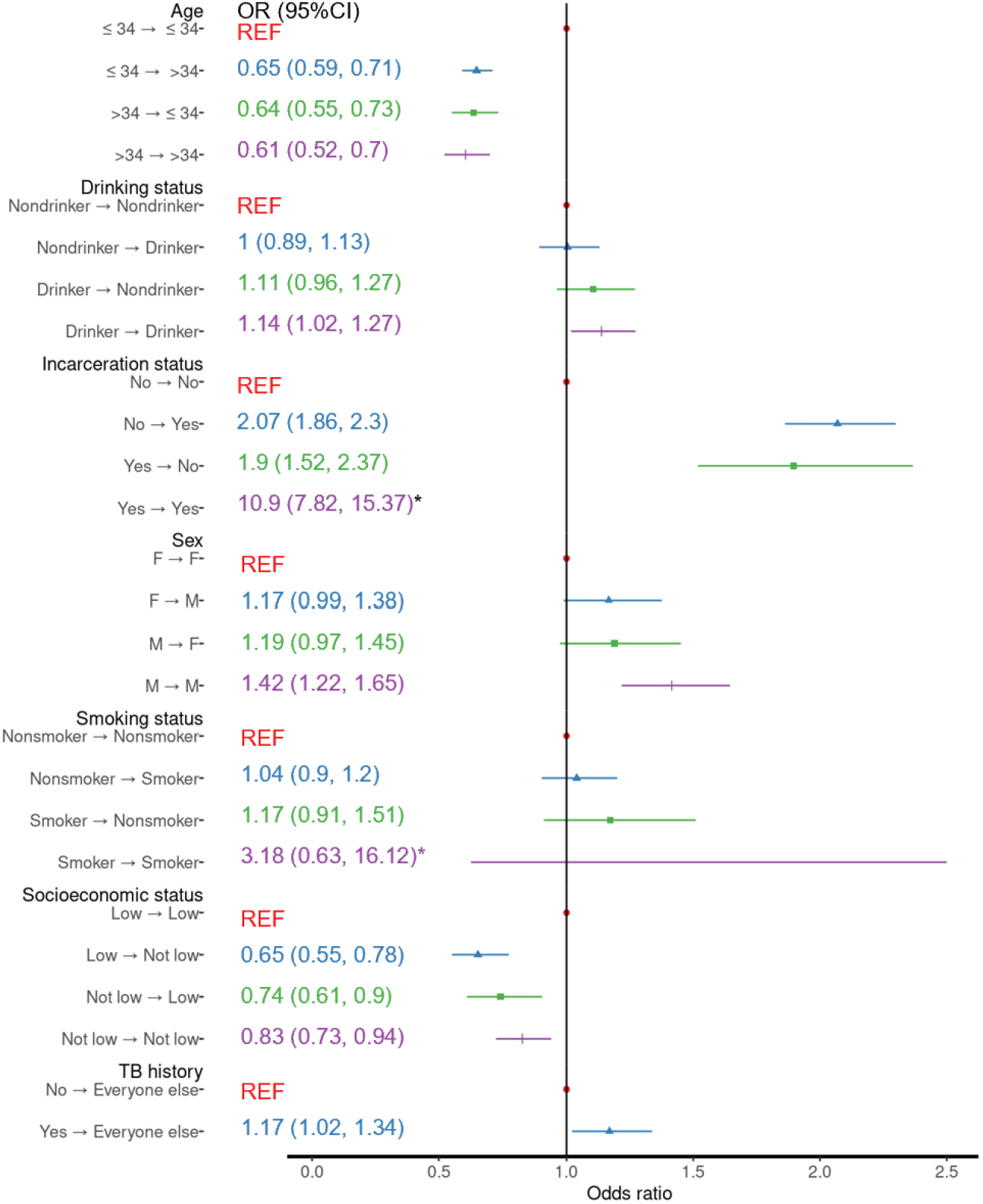
Adjusted naïve Bayes modified close genetic relatedness odds ratios 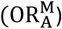 and 95% confidence intervals for covariate contributions from the Lima, Peru cohort dataset. Variables included in this analysis had unadjusted naïve Bayes modified close genetic relationship odds ratios with p-value ≥ 0.1. *The value is out of scale and has not been included or entirely represented.

## 4. DISCUSSION

We describe a method to generate adjusted ORs describing the relationship between pathogen genetic relatedness and pair level covariates using bootstrapping within mlTransEpi.^17^ Importantly, this approach accounts for correlation in the data, which has been lacking in other approaches.^14, 15, 16, 24^ We show using data from a cohort study in Lima, Peru that our method estimates similar point estimates compared to logistic regressions but with increased standard errors, reflecting correlation adjustments.

The iterative process accounts for correlation that arises in pathogen genetic relatedness datasets from two factors: (1) a SNP distance-based cut-off can yield multiple potential infectors for one infectee and (2) one infector can appear in multiple potential transmission instances. mlTransEpi corrects for the first source by randomly choosing one ‘true’ infector per infectee per iteration, removing multi-potential-infector correlation. The second is addressed by bootstrapping the logistic regression model within each fold and iteration, as bootstrapped standard errors are robust to correlation.^18^

Our findings largely agree with those from the Trevisi et al. analysis.^14^ Both show that transmission is more likely in pairs with at least one male.^14^ Men are well-documented as over-represented in the TB epidemic,^2, 19^ possibly due to lower rates of care seeking^25, 26^ and/or greater sputum expectoration.^27^ We also echo the finding that the social risk factors (smoking, drinking, incarceration) increase transmission odds, consistent with other studies.^28, 29, 30, 31, 32^ We no longer show a significant increase in transmission smoker-smoker transmission pairs, although this could be due to the low incidence of such pairs.

Both methods find decreased odds of transmission when the infector is ≥34. However, mlTransEpi also finds significantly decreased odds of transmission when the infector is <34 and the infectee is ≥34. Younger patients have higher TB rates,^3, 19, 33^ hypothesized to be due to more social contacts and thus greater transmission contribution, though evidence is limited.^34, 35^ Two studies in the Netherlands support younger adults as more likely infectors, showing 67% of source cases from ages 15-34, versus 58% of secondary cases.^36, 37^ However, as these percentages are not population adjusted, they may simply be reflective of the high rates of TB in these populations.

We compare mlTransEpi to GenePair,^6^ an existing method which employs a different approach to answer a similar question. GenePair uses transmission probabilities generated by TransPhylo, a method that infers transmission trees using WGS and observation dates,^10^ to generate ORs from a zero-inflated distribution model with individual-level spatially correlated random effect parameters to account for correlation. The model includes a binary component to account for the large proportion of transmission probabilities of 0. Thus, their ORs describe the odds of having non-zero transmission probabilities. GenePair employs many strong assumptions, including the accuracy of the transmission probabilities from TransPhylo. Additional work has shown that using TransPhylo to inform logistic regression biases ORs towards the null as the fraction of sampled cases fed into TransPhylo decreases,^13^ implying that GenePair likely works best in low burden or high sampling coverage setting.

Our method, in comparison, estimates an OR describing the relationship between pathogen genetic relatedness and pair level covariates, a demonstrated proxy for transmission-covariate relationships. We do not assume full transmission network sampling, nor that an infectee’s true infector is in the dataset. Instead, we create possible transmission pairs based on timing, designating potential links and non-links by SNP distances.

We also avoid assuming SNP distance thresholds firmly define links/non-links, as we randomly select one true infector per iteration. Sensitivity analyses show our method is robust to varying SNP distance thresholds (Supplementary Materials Figure S4). That said, future work aims to enhance link selection with additional WGS data and other data such as geographical distances.

In this work, we assume directionality of potential transmission pairs from the time of diagnosis. We know that care seeking delays exist for a myriad of reasons,^38, 39, 40^ as such it is possible that the individual diagnosed second actually infected the first. However, a sensitivity analysis where we allowing ‘negative’ time (infectee diagnosed before infector) showed ORs remained constant despite varying time windows for infector/infectee pair specification, except for low-incidence pairs (e.g., both smokers or incarcerated history) where window changes significantly altered pair instances (Supplemental Materials Figure S6).

We produce adjusted ORs via bootstrapped logistic regression within an iterative algorithm, appropriately accounting for pairwise genetic relatedness data correlation. This method is applicable to any disease with WGS to estimate covariate-transmission associations, furthering knowledge of transmission risk factors. Understanding these risk factors is crucial for better transmission control and outbreak prevention.

## Data Availability

Data cannot be shared publicly to protect study participant privacy. Code is available upon request.

## Notes

### Competing Interest Statement

The authors have declared no competing interest.

### Funding Statement

The authors disclosed receipt of the following financial support for the research, authorship, and/or publication of this article: ANS is funded by the National Institute of Allergy and Infectious Disease, National Institutes of Health (grant number 1F31AI183782-01A1). MBB is funded by the National Institutes of Health and the National Institute of Allergy and Infectious Diseases grants (grant number K01AI151083). MBM is funded by funded by the National Institutes of Health and the National Institute of Allergy and Infectious Diseases grants (grant numbers U01AI057786, U19AI076217, U19AI109755, U19AI111224, and U19AI142793). LFW is funded by the National Institutes of Health (grant number R35GM141821). The content of the article is solely the responsibility of the authors and does not necessarily represent the views of the funding agencies. The funders had no role in the decision to publish this manuscript.

### Author Declarations

The Harvard School of Public Health institutional review board and the Research Ethics Committee of the National Institutes of Health of Peru gave ethical approval for this work. All study participants provided voluntary written informed consent prior to study participation.

