## Supplemental Materials for "An updated method to estimate factors associated with tuberculosis transmission using whole genome sequencing and other additional metadata"

**Table of Contents**

**S1. Iterative algorithm**.….………………………………………………………………………………………2

**S1. Bootstrapping**…………………………………………………………………………………………………3

**S2. Simulation studies**…………………………………………………………………………………………..5

**Section S1. Iterative algorithm**

The following algorithm describes the iterative estimation procedure shown in Main Text Figure 1 used to estimate the adjusted odds ratios within mlTransEpi.

1. Create a dataset of all possible infector-infectee pairs from individuals for whom information to define probable links is available.
2. Divide dataset from 1. into Training, consisting of pairs determined to be high probability transmission links or non-links, and Prediction, consisting of all other pairs.
3. In the Training data from step 2:
   a. For each infectee, randomly select exactly one of its potential infectors to designate as "true links" (L_ij_ = 1).
   b. Designate all remaining pairs as not linked (L_ij_ = 0).
4. Split Training data into *n* folds.
   a. Reserve one fold and add it into the Prediction dataset.
   b. Set the predicted probabilities for training set pairs to 1 for links (L_ij_ = 1) and 0 for non-links (L_ij_ = 0).
   c. For any infectee that has a designated true link (L_ij_ =1) from Step 3, move all other pairs with this infectee in the Prediction into the Training set and designate as non-links (L_ij_ = 0).
   d. Use the training set to estimate the adjusted odds ratios via bootstrapped logistic regression.
   e. Repeat steps a.-d. *n* times to allow each fold a turn in the Prediction set.
5. Repeat steps 3 and 4 to allow for multiple designations of “true links.”
6. Combine estimates using Rubin’s rules to obtain final pooled estimates.

**Section 2. Bootstrapping**

For the simulation study, we perform a true bootstrap of the logistic regression within each iteration and cross-validation fold using the following method:

1. Perform a bootstrap resampling from the $n_{1}$ probable linked observations.
2. Perform a bootstrap resampling from the $n_{0}$ probable not-linked observations.
3. Perform a multivariate logistic regression analysis on the sample from steps 1 and 2, and save logistic regression coefficients.
4. Repeat steps 1 and 2 $r$ times, producing $r$ sets of bootstrap regression coefficients
5. Take the mean and standard deviation of the $r$ regression coefficients for each variable in the model to be the bootstrapped estimate and standard deviation for a cross-validation fold within an iteration.

In a model with $m$ iterations and $n$ cross validation folds, the iterative estimation procedure results $m*n$ estimates of each odds ratio. We pool these $m*n$ for each odds ratio using Rubin’s Rules^1^ to obtain the final standard errors for each odds ratio.

Rather than perform a true bootstrap, where each sample is the size of the dataset, we employed a modified sampling scheme motivated by Trevisi et al.^2^ As performing a full bootstrap within the iterative estimation procedure is too computationally intensive, we modify the sampling procedure. Instead, we follow the below algorithm:

1. Perform a bootstrap resampling from the $n_{1}$ probable linked observations.
2. Take a subsample of size $m$ from the $n_{0}$ probable not linked observations and perform a bootstrap resampling on those $m$ (i.e. sample $m$ cases with replacement from the subsample).
3. Perform a multivariate logistic regression analysis on the sample from steps 1 and 2 and save logistic regression coefficients.
4. Repeat steps 1 and 2 $r$ times, producing $r$ sets of bootstrap regression coefficients
5. Take the mean and standard deviation of the $r$ regression coefficients for each variable in the model to be the bootstrapped estimate and standard deviation for a cross-validation fold within an iteration.

We can show via simulation that excluding non-linked cases does not impact estimates. We simulate 100 TB outbreaks of minimum size 400 and calculate bootstrapped odds ratios and standard errors using all cases, 40% of cases, and 70% of cases. Both odds ratios and their standard errors are consistent across all proportions of non-links sampled (Figure S1). We also performed sensitivity analyses using a single logistic regression (not within the iterative algorithm) using the cohort data from Lima, Peru. We varied $m$ between 5,000 and the full sample size (3,069,784). We performed this sensitivity analysis on only one logistic regression model, as opposed to within the iterative scheme, as only minor changes are made to the data structure within each iteration and thus we expect results to hold. Results are shown in Figure S2.

We can also think of our pairwise dataset as a case control study, with linked pairs being the cases and non-linked pairs being controls. In a pairwise dataset generated from $N$ cases, we expect probable links to be of the $N$ (each case has only one true link after modification within each iteration) and non-links to be on the order of $\frac{N^{2}}{2}$ (all other pairwise combinations in which the infector was documented before the infectee). It has been shown that while variance decreases with additional controls for each case, gains in precision are minimal after five controls per case.^3^

Figure S1. Bootstrapped odds ratios standard errors intervals for simulated outbreaks with varying sampled non-link proportions.

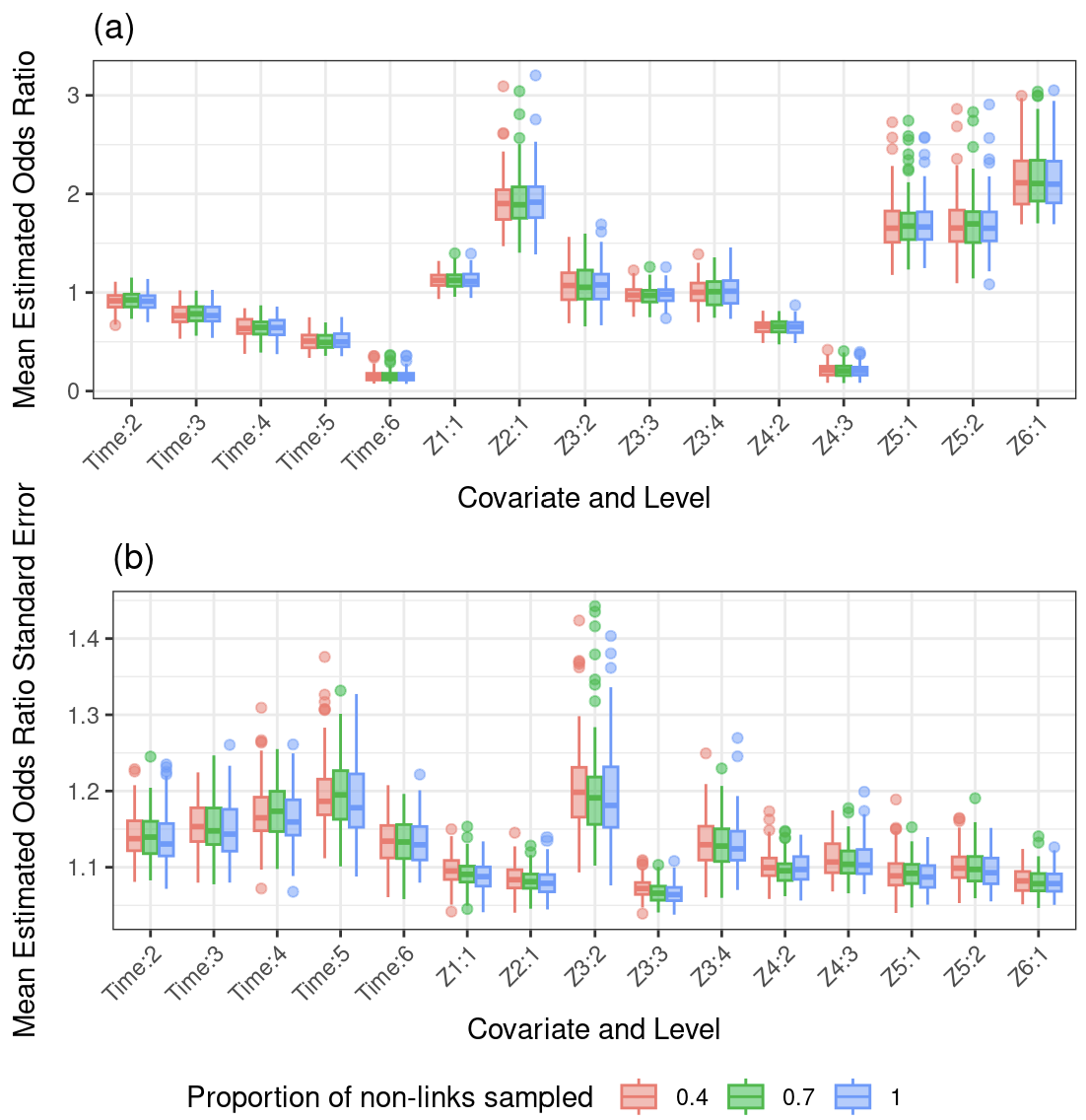

**Figure S2**. Odds ratios and 95% confidence intervals for adjusted logistic regression model using Lima, Peru data with varying bootstrap sample sizes.

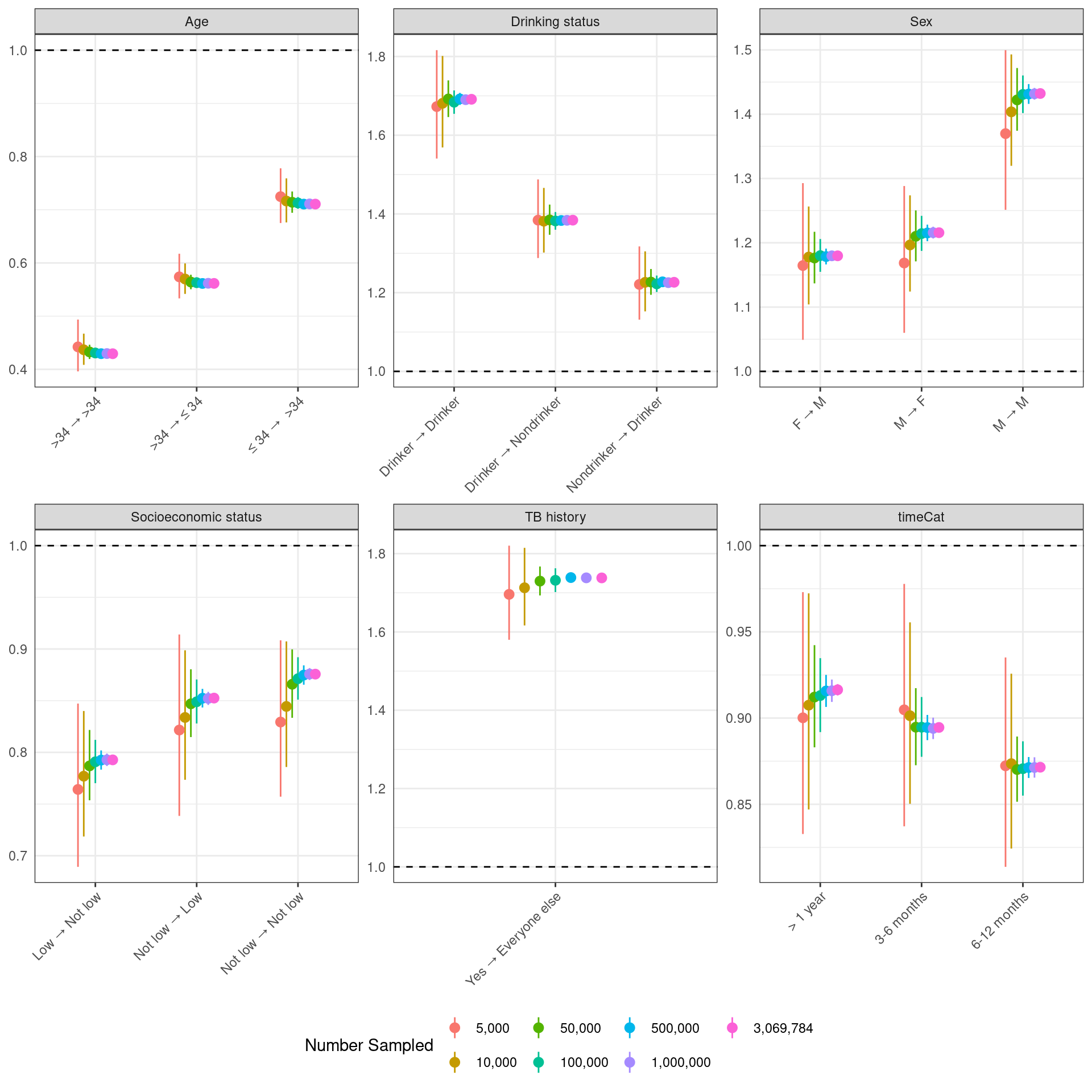

**Section 2. Simulation study**

**Simulation details**

The R package TransPhylo simulates disease outbreak using a negative binomial distribution to specify the reproductive number and a gamma distribution to describe the serial interval (the time between onset of symptoms between two cases). The simulated outbreak will continue until it (a) dies out, (b) runs for the specified period of time, or (c) reaches the specified size. TransPhylo also allows for simulated sampling times (the time that an individual was diagnosed) in addition to simulated infection times through an additional gamma distribution that represents the time in between infection and diagnosis. The R package phangorn takes the simulated transmission trees simulates a genetic sequence using a specified mutation rate. This simulation scheme was originally described in Stimson et al.^4^ and extended by Leavitt et al.^5^ to include multiple transmission chains.

We specify our simulation as in Leavitt et al.^5^ For an outbreak, we simulate multiple transmission chains with a minimum of 2 cases each and allow them to run for 20 years. Chains are simulated iteratively and independently until the size of the outbreak exceeds 300. The genetic sequences are simulated as a random sequence of 300 base pairs. As each chain within an outbreak has different phylogenetic trees, we expect each the genetic sequences between individuals in separate transmission chains to differ by approximately 300 single nucleotide polymorphisms (SNPs).

We assume the reproductive number to have a negative binomial distribution with number of successes (*r)* equal to 1 and success probability (*p*) of 0.5. ^6^ We assume the generation interval and sampling interval to both be gamma distributed with shape parameter (α) of 1.2 and scale (θ) of 2, shifted by 0.25 such that infection events are separated by at least 3 months.^6^ We assume a mutation rate of 0.5 SNPs per genome per year.^7^

We simulate 6 covariates at the individual level, $X_{i}, i=1, \ldots, 6$, using the outbreak’s phylogenetic tree. We first simulate covariates values of $X_{i}$for each source case according to the values specified in the second column of Table S1. We then looped through the individuals and assigned covariate values based on the value of their infector, as specified in the third column of Table S1. We then created pair level variables, $Z_{i}, i=1, \ldots, 6$, as specified in the fourth column of Table S1. To better understand the behavior of the adjusted odds ratio, we simulated $X_{1}$ and $X_{5}$ to be correlated. For the source cases, $X_{1}$ and $X_{5}$ were simulated as correlated binary variables with a shared success probability of 0.6 and a correlation of 0.8. While looping through the remaining cases to assign values based off the infector, covariates $X_{1}$ and $X_{5}$ were again simulated from binary variables with a correlation of 0.8.

**Table S1**. Simulated covariates structures. $X_{i}$ refers to the individual level covariate and $Z_{i}$ refers to the pair level covariate. Source variable frequency refers to the frequency for the starts of transmission chains. Linked variable frequency refers to the frequency for all other individuals. Covariates $X_{1}$ and $X_{5}$ were simulated as correlated binary variables with correlation of 0.8.

| **Variable** | **Source individual level variable frequency** | **Linked individual level variable frequency** | **Paired variable construction** | **Motivating Example** |
| --- | --- | --- | --- | --- |
| $X_{1}/Z_{1}$ | 0: 60%; 1: 40% | *If* $X_{1}=X_{5}=0$*:*  80% chance of same  20% chance of different  *If* $X_{1}=X_{5}=1$*:*  20% chance of same  80% chance of different  *If* $X_{1}\neq X_{5}$*:*  50% chance of same  50% chance of different | $Z_{1}=1$ if same  $Z_{1}=0$ if different | Sex |
| $X_{2}/Z_{2}$ | a: 50%; b: 30%;  c: 15%; d:5% | 70% chance of same  30% chance of different | $Z_{2}=1$ if same  $Z_{2}=0$ if different | Nationality |
| $X_{3}/Z_{3}$ | 0: 70%; 1: 30% | *Infector is a:*  80% of a-a; 20% of a-b  *Infector is b:*  70% of b-b; 30% of b-a | $Z_{3}=1$ if 0-0  $Z_{3}=2$ if 1-1  $Z_{3}=3$ if 0-1  $Z_{3}=4$ if 1-0 | Age group |
| $X_{4}/Z_{4}$ | a: 5%; b: 5%;  c: 5%; d:20%;  e: 30%; f:10%;  g:10%; h: 5%;  i:5%; j:5% | 60% chance of same  35% chance of neighbors  5% chance of other | $Z_{4}=1$ if same  $Z_{4}=2$ if neighbors  $Z_{4}=3$ otherwise | Country of origin |
| $X_{5}/Z_{5}$ | 0: 60%; 1: 40% | *If* $X_{1}=X_{5}=0$*:*  80% chance of same  20% chance of different  *If* $X_{1}=X_{5}=1$*:*  20% chance of same  80% chance of different  *If* $X_{1}\neq X_{5}$*:*  50% chance of same  50% chance of different | $Z_{5}=1$ if 0-0  $Z_{5}=2$ if 1-1  $Z_{5}=3$ if different | Social risk factor |
| $X_{6}/Z_{6}$ | a: 34%; b: 34%;  c: 33% | 75% chance of same  25% chance of different | $Z_{6}=1$ if same  $Z_{6}=0$ if different | Common location |
| Time | -- | -- | Time = 1 if <1 year  Time = 2 if 1-2 years  Time = 3 if 2-3 years  Time = 4 if 3-4 years  Time = 5 if 4-5 years  Time = 6 if >5 years | -- |

**Figure S3**. Mean estimated odds ratios from 100 simulated outbreaks describing the unadjusted association between covariates and transmission (OR^T^), adjusted association between covariates and transmission $(\mathrm{OR}_{A}^{T}$), unadjusted odds ratios describing the association between covariates and iteratively modified close genetic relatedness (OR^M^), and adjusted bootstrapped odds ratios describing the association between covariates and iteratively modified close genetic relatedness $(\mathrm{OR}_{A}^{M}$). Row (a) shows results across varying generation interval distribution scale parameters, row (b) shows results across varying sampling distribution scale parameters, and row (c) shows results varying reproductive numbers.

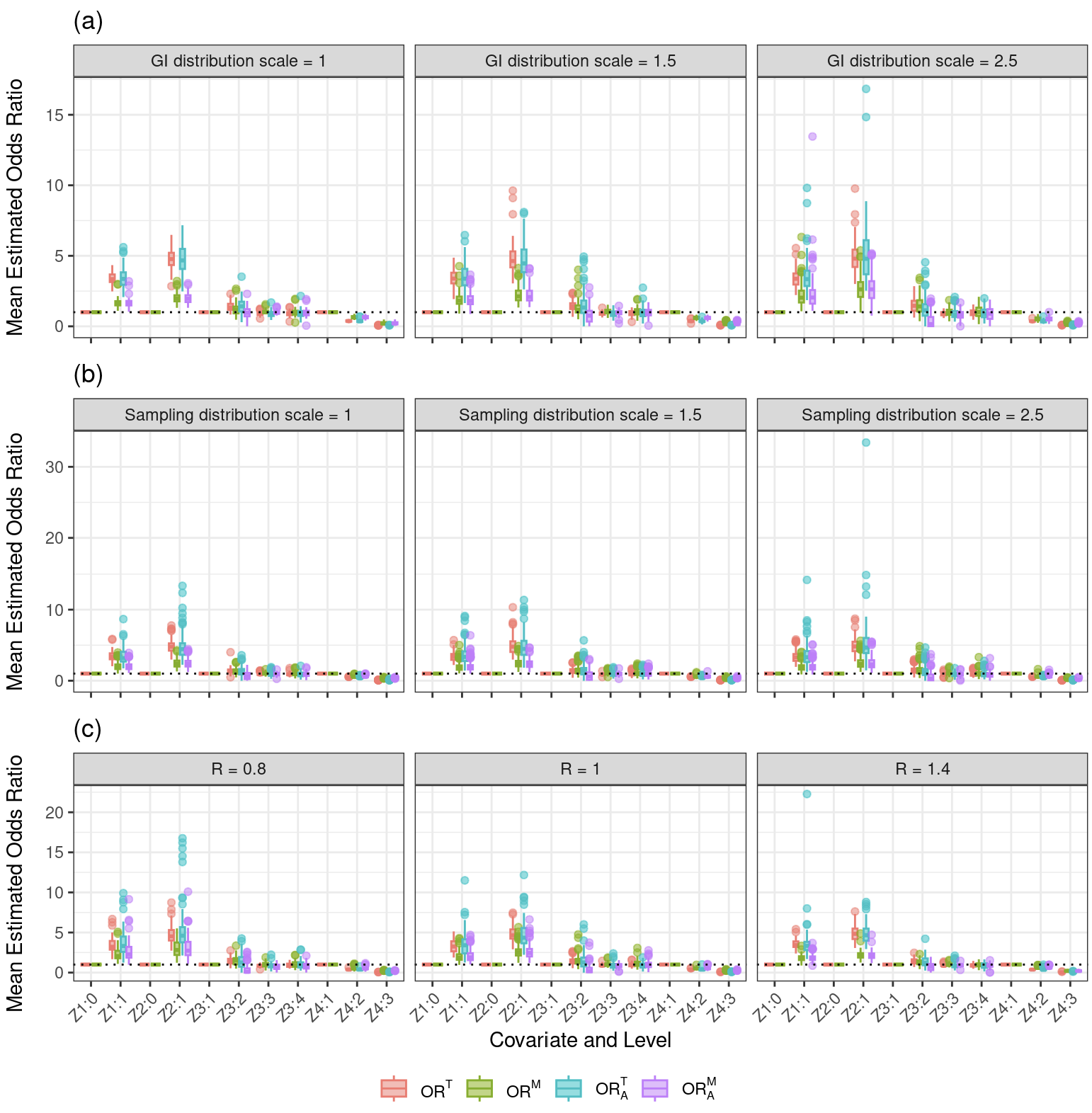

**Figure S4**. Mean estimated odds ratios from 100 simulated outbreaks describing the unadjusted association between covariates and transmission (OR^T^), adjusted association between covariates and transmission $(\mathrm{OR}_{A}^{T}$), unadjusted odds ratios describing the association between covariates and iteratively modified close genetic relatedness (OR^M^), and adjusted bootstrapped odds ratios describing the association between covariates and iteratively modified close genetic relatedness $(\mathrm{OR}_{A}^{M}$), stratified by single nucleotide polymorphisms (SNP) difference.

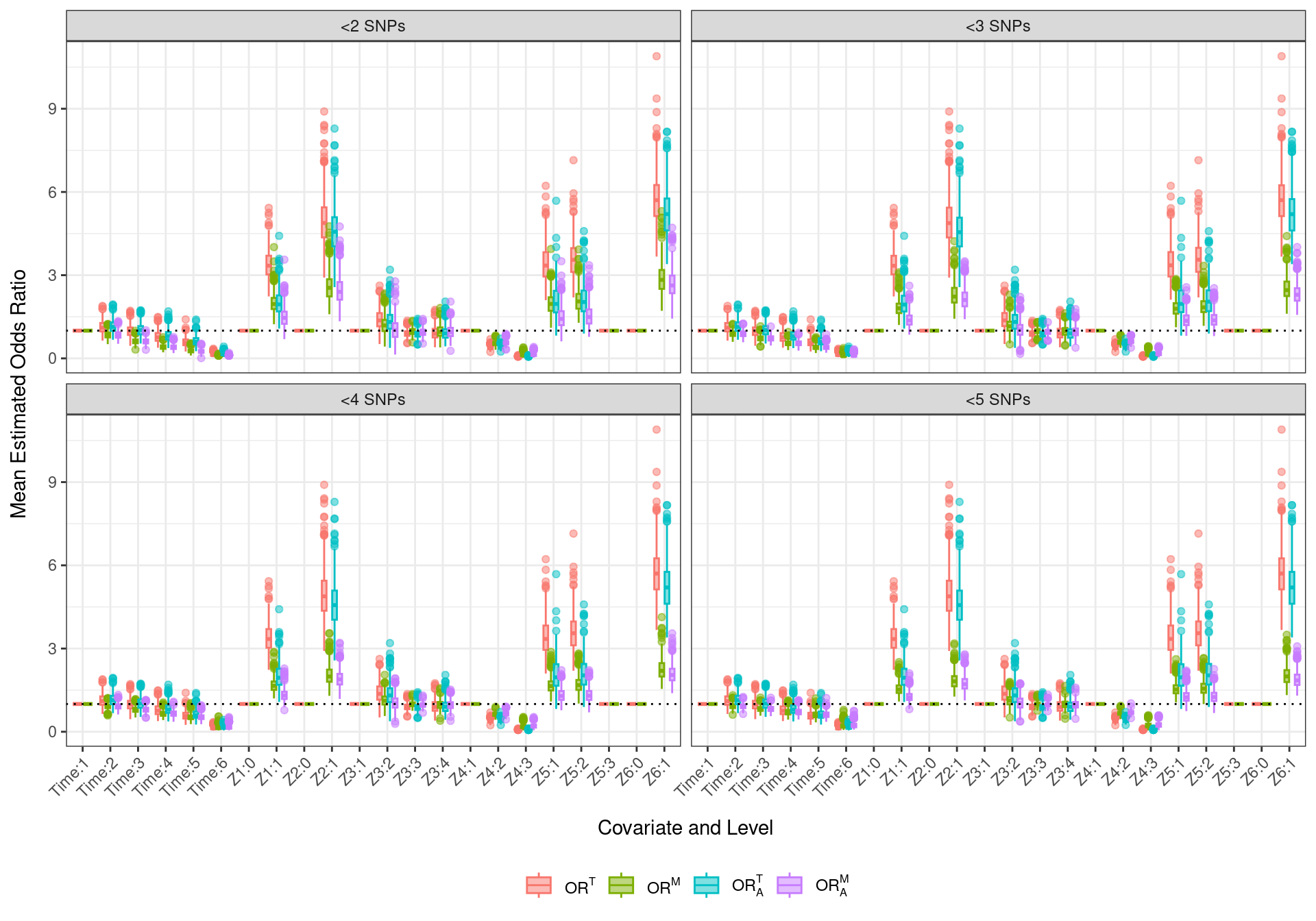

**Figure S5**. Error and confidence interval (CI) coverage across 100 simulated outbreaks with independent covariates. The odds ratio describing the association between transmission and the covariates (OR^T^) is considered truth for the odds ratio describing the association between covariates and iteratively modified close genetic relatedness (OR^M^) and the adjusted odds ratio describing the association between transmission and covariates ($\mathrm{OR}_{A}^{T})$ is considered truth for the adjusted bootstrapped odds ratios describing the association between covariates and iteratively modified close genetic relatedness $(\mathrm{OR}_{A}^{M}$) and the adjusted odds ratios describing the association between covariates and iteratively modified close genetic relatedness $(\mathrm{OR}_{A}^{M}'$) in the calculation of mean absolute percentage error (MAPE), mean standard error (MSE), and conference interval coverage.

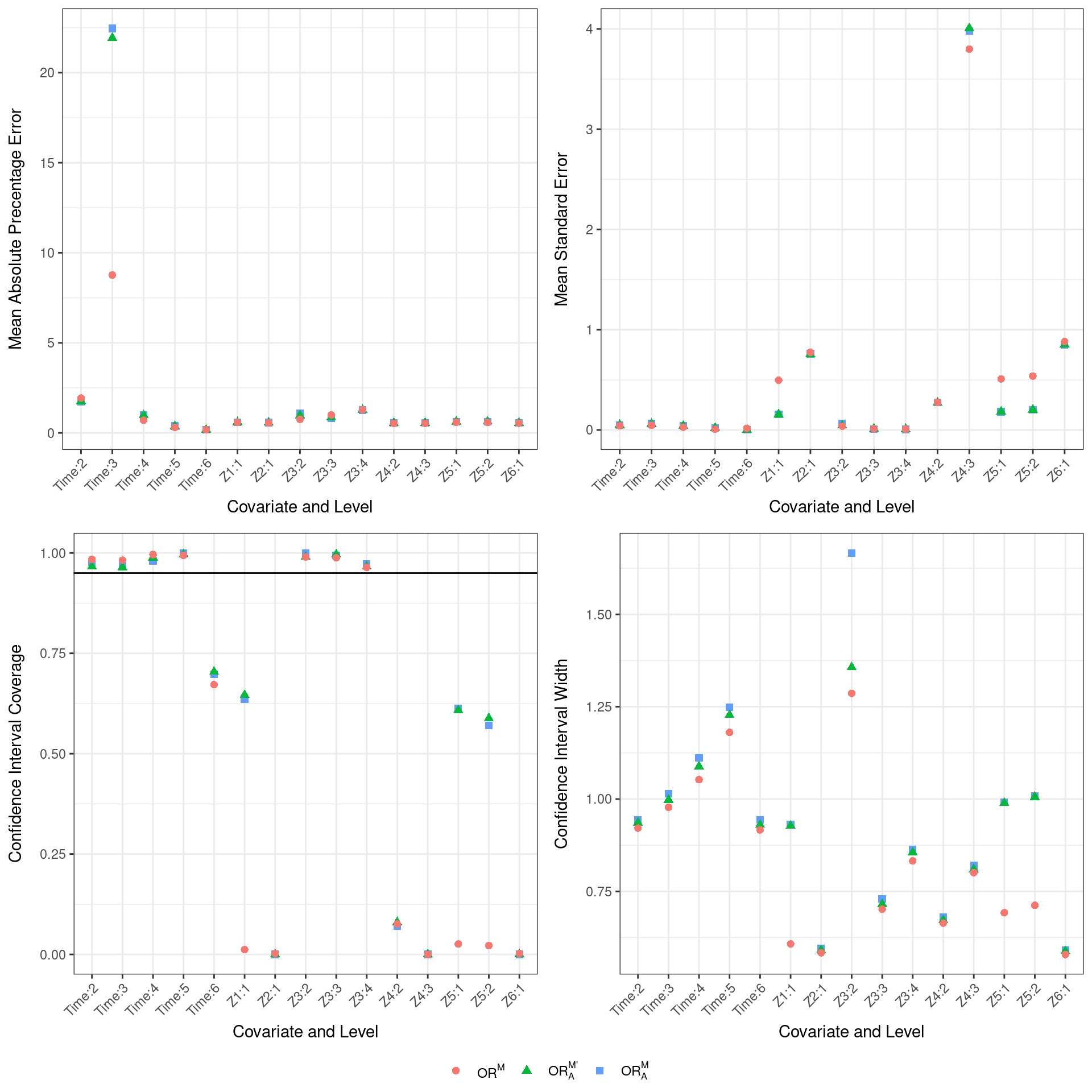

**Table S2**. Unadjusted naïve Bayes modified close genetic relatedness odds ratios (OR^M^) and 95% confidence intervals for covariate contributions from the Lima, Peru cohort dataset.

| Variable | Category | Unadjusted OR (95% CI) |
| --- | --- | --- |
| Age | ≤ 34 → ≤ 34 | REF |
|  | ≤ 34 → >34 | 0.66(0.52, 0.83) |
|  | >34 → ≤ 34 | 0.65 (0.5, 0.84) |
|  | >34 → >34 | 0.6 (0.44, 0.83) |
| Cavitation | Yes → Everyone else | 1.02 (0.83, 1.26) |
|  | No → Everyone else | REF |
| Cough > 1 month | Yes → Everyone else | 0.93 (0.76, 1.13) |
|  | No → Everyone else | REF |
| Diabetes mellitus | No → No | REF |
|  | No → Yes | 0.66 (0.42, 1.04) |
|  | Yes → Everyone else | 0.7 (0.43, 1.13) |
| Drinking status | Nondrinker → Nondrinker | REF |
|  | Nondrinker → Drinker | 1.16 (0.89, 1.5) |
|  | Drinker → Nondrinker | 1.26 (0.97, 1.63) |
|  | Drinker → Drinker | 1.57 (1.23, 2) |
| Incarceration status | No → No | REF |
|  | No → Yes | 2.31 (1.71, 3.12) |
|  | Yes → No | 2.29 (1.6, 3.27) |
|  | Yes → Yes | 14.87 (8.43, 26.21) |
| HIV status | No → No | REF |
|  | No → Yes | 1.48 (0.99, 2.21) |
|  | Yes → No | 1.52 (1, 2.32) |
|  | Yes → Yes | 5.84 (1.92, 17.74) |
| Public transportation  use | Light → Light | REF |
|  | Light → Heavy | 1.02 (0.8, 1.31) |
|  | Heavy → Light | 1.12 (0.87, 1.45) |
|  | Heavy → Heavy | 1.13 (0.88, 1.45) |
| Sex | F → F | REF |
|  | F → M | 1.22 (0.85, 1.75) |
|  | M → F | 1.31 (0.9, 1.89) |
|  | M → M | 1.74 (1.27, 2.39) |
| Smear status | Positive → Everyone else | 1.05 (0.84, 1.31) |
|  | Negative → Everyone else | REF |
| Smoking status | Nonsmoker → Nonsmoker | REF |
|  | Nonsmoker → Smoker | 1.39 (0.87, 2.22) |
|  | Smoker → Nonsmoker | 1.81 (1.13, 2.89) |
|  | Smoker → Smoker | 9.32 (2.81, 30.92) |
| Socioeconomic status | Low → Low | REF |
|  | Low → Not low | 0.64 (0.46, 0.89) |
|  | Not low → Low | 0.74 (0.52, 1.04) |
|  | Not low → Not low | 0.8 (0.6, 1.05) |
| TB history | Yes → Everyone else | 1.28 (1, 1.64) |
|  | No → Everyone else | REF |
| Time category | <3 months | REF |
|  | 3-6 months | 0.77 (0.59, 0.99) |
|  | 6-12 months | 0.66 (0.52, 0.84) |
|  | > 1 year | 0.52 (0.38, 0.73) |
| Work outside the house | No → No | REF |
|  | No → Yes | 0.9 (0.71, 1.14) |
|  | Yes → No | 0.98 (0.76, 1.25) |
|  | Yes → Yes | 0.92 (0.69, 1.23) |

**Figure S6.** Estimated adjusted naïve Bayes modified close genetic relatedness odds ratios $(\mathrm{OR}_{A}^{M}$) and 95% confidence for the Lima, Peru cohort dataset. We vary the time between diagnosis of the infector and infectee. Positive time indicates that the infector was only allowed to be diagnosed before the infectee and negative time indicates the infector was allowed diagnosed after the infectee.

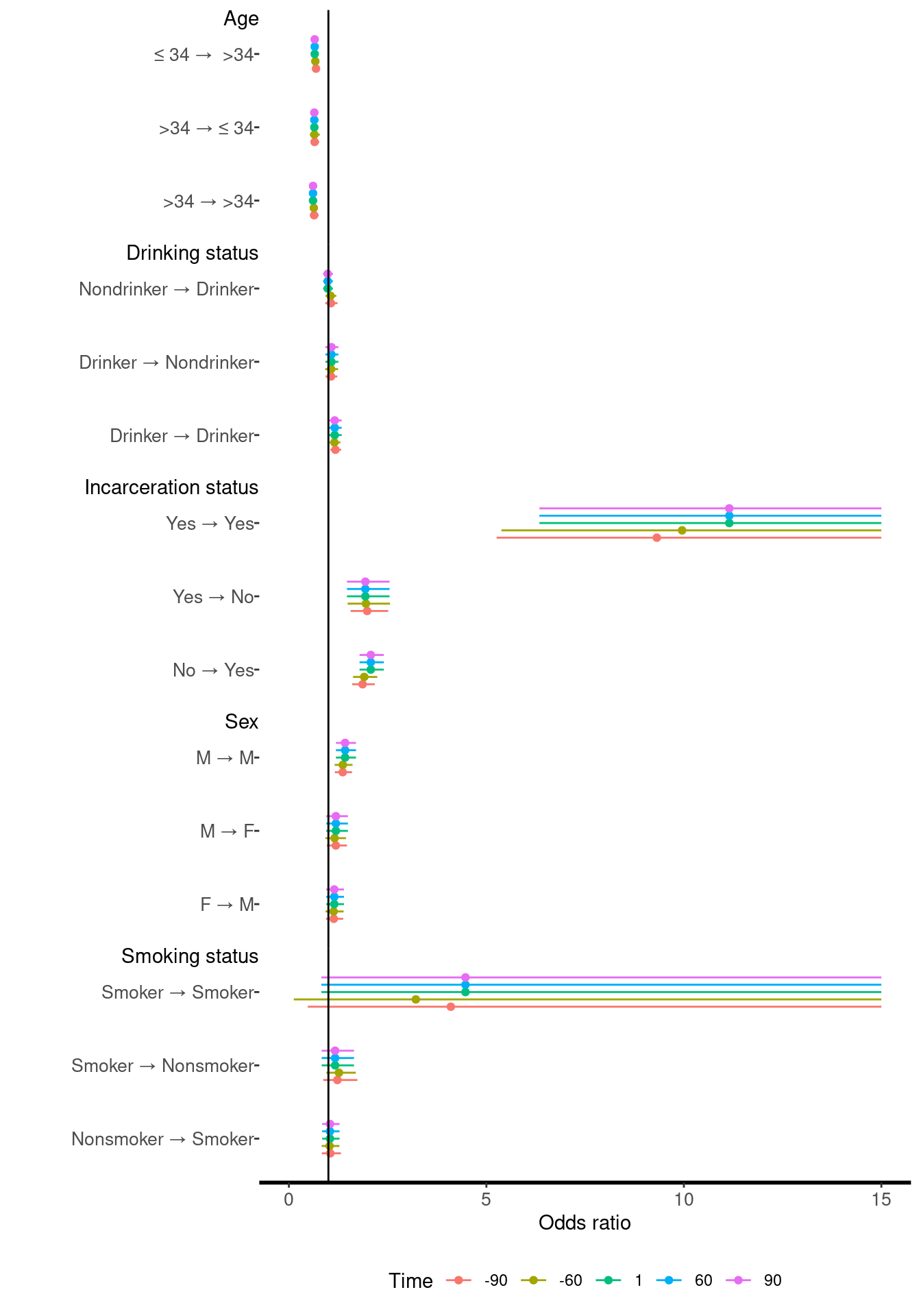
